# Wildland Fire-Related Smoke PM_2.5_ and Cardiovascular Disease ED Visits in the Western United States

**DOI:** 10.1101/2024.10.08.24314367

**Authors:** Linzi Li, Wenhao Wang, Howard H. Chang, Alvaro Alonso, Yang Liu

## Abstract

**Background:** The impact of short-term exposure to fine particulate matter (PM_2.5_) due to wildland fire smoke on the risk of cardiovascular disease (CVD) remains unclear. We investigated the association between short-term exposure to wildfire smoke PM_2.5_ and Emergency Department (ED) visits for acute CVD in the Western United States from 2007 to 2018.

**Methods:** ED visits for primary or secondary diagnoses of atrial fibrillation (AF), acute myocardial infarction (AMI), heart failure (HF), stroke, and total CVD were obtained from hospital associations or state health departments in California, Arizona, Nevada, Oregon, and Utah. ED visits included those that were subsequently hospitalized. Daily smoke, non-smoke, and total PM_2.5_ were estimated using a satellite-driven multi-stage model with a high resolution of 1 km. The data were aggregated to the zip code level and a case-crossover study design was employed. Temperature, relative humidity, and day of the year were included as covariates.

**Results:** We analyzed 49,759,958 ED visits for primary or secondary CVD diagnoses, which included 6,808,839 (13.7%) AFs, 1,222,053 (2.5%) AMIs, 7,194,474 (14.5%) HFs, and 808,396 (1.6%) strokes. Over the study period from 2007-01-01 to 2018-12-31, the mean smoke PM_2.5_ was 1.27 (Q1: 0, Q3: 1.29) µg/m^3^. A 10 µg/m^3^ increase in smoke PM_2.5_ was associated with a minuscule decreased risk for AF (OR 0.994, 95% CI 0.991-0.997), HF (OR 0.995, 95% CI 0.992-0.998), and CVD (OR 0.9997, 95% CI 0.996-0.998), but not for AMI and stroke. Adjusting for non-smoke PM_2.5_ did not alter these associations. A 10 µg/m^3^ increase in total PM_2.5_ was linked to a small increased risk for all outcomes except stroke (OR for CVD 1.006, 95% CI 1.006-1.007). Associations were similar across sex and age groups.

**Conclusion:** We identified an unexpected slight lower risk of CVD ED visits associated with short-term wildfire smoke PM_2.5_ exposure. Whether these findings are due to methodological issues, behavioral changes, or other factors requires further investigation.

## Background

The surge in wildfire occurrences globally in recent decades has raised alarms about the role of climate change.^1^ The repercussions of climate change include exacerbating fire frequency and endangering human livelihoods, ecosystems, and public health.^1–3^ Coupled with warming and drought climatic conditions, severe fire events can catalyze vegetation changes, implicating rapid ecosystem shifts and reductions in valued resources.^1,4,5^ In the United States, the wildfire-burned area has increased approximately fourfold over the past four decades, and the trend is expected to continue as a result of climate warming, earlier spring, and other factors.^6^ The western US, an area prone to wildfire disasters, often faces consecutive years marked by both drought and heightened fire weather.^7^ Since 1986, the frequency of large wildfires and the area burned in the western US have been observed to be four and six and a half times the average level between 1970 and 1986.^6^ Smoke from wildfires significantly contributes to fine particulate matter in the atmosphere, a form of air pollution consisting of particles with a diameter of less than 2.5 micrometers (PM_2.5_).^8,9^ The contribution of wildfire smoke to PM_2.5_ concentrations across the US has increased since the mid-2000s, now representing up to half of the total PM_2.5_ in western regions, a stark increase from less than 20% around 2015.^6,10^ Additionally, smoke influences the trends in extreme PM_2.5_ days in western and mid-western states. Evidence shows that since 2012, the rise in the number of days above 35 µg/m^3^ of PM_2.5_ would have been lower without wildfire smoke, and the days surpassing 35 µg/m^3^ of PM_2.5_ would not have exceeded the threshold absent wildfire smoke on that day.^11^

Long-term exposure to ambient PM_2.5_ is linked with several adverse cardiometabolic health outcomes and cardiopulmonary disease, including increased blood glucose, endothelial dysfunction, incident cardiovascular disease (CVD) events, chronic obstructive pulmonary disease, pneumonia, and all-cause mortality.^12–14^ Recognized as a substantial contributor to ambient PM_2.5_ levels, wildfire smoke has been identified as a potential environmental risk factor for CVD, which includes a range of heart and blood vessel disorders and stroke.^9,15–18^ Between 2017 and 2020, nearly 10% of US adults, totaling 28.6 million individuals, experienced some form of CVD, including coronary artery disease (CAD), heart failure (HF), stroke.^19^ Moreover, CVD complications significantly compound the burden for both individuals and the healthcare system worldwide. CVD can exacerbate or increase the risk of other chronic conditions, such as diabetes, kidney disease, and functional decline, further impairing the quality of life and potentially leading to early death.^19^ Globally, CVD is the primary cause of death, responsible for 19.9 million (95% UI 18.4, 21.2) deaths in 2021 with 13.3% of these CV deaths attributable to ambient PM_2.5_ exposure.^20,21^ In the US, among the Medicare enrollees, exposure to a 12-month chronic PM_2.5_ was associated with 1.56-fold (95% CI 1.55, 1.57) increase in CV mortality.^13^

Previous literature has documented the potential pathophysiologic mechanisms through which wildfire smoke can contribute to CVD. Inhaled PM that accumulates in the alveoli may be a trigger of a series of adverse reactions that lead to CVDs, including inflammation, oxidative stress, thrombosis and coagulation, and vascular dysfunction.^22–24^ Individual susceptibility conditions and concomitant existing air co-pollutants, such as preexisting CAD and ozone could worsen these effects.^22,25^ Numerous epidemiological studies have investigated the cardiovascular effects of wildfire smoke. However, the transient nature of wildfire events introduces challenges in defining the exposure time window and ensuring consistency in the measurement of PM during wildfire events. There are discrepancies in whether the acute PM exposure should be based on 3-day averages or one-hour daily peak averages, and whether chronic PM exposure should be quantified using annual averages or extreme daily wildfire PM. Although certain past studies have indicated that wildfire smoke PM_2.5_ increased CVD morbidity and mortality, these associations have not been consistent across regions, populations, and periods, partially due to different wildfire smoke PM_2.5_ concentration measures.^26,27^ Measuring smoke PM_2.5_ have challenges, including limited spatial monitoring coverage, Aerosol Optical Depth (AOD) data availability, model performance in extreme concentrations, temporal and spatial variability and etc. In a global time series study in 749 locations, short-term wildfire smoke PM_2.5_ did not have a substantial impact on CV mortality in the US (RR 1.014, 95% CI 0.998, 1.031), while wildfire-related PM_2.5_ was associated with a higher risk of CV mortality in the pooled result of all the areas.^28^

Therefore, we aim to add to the evidence by exploring the association between exposure to short-term wildfire smoke PM_2.5_ exposure and emergency department (ED) visits for CVD in the western United States from 2007 to 2018, using satellite-driven exposure measures.

## Methods

### Emergency department visit data

ED visits for CVDs in five western US states were obtained from hospital associations or state health departments. The states and corresponding data ranges are: California (2007-2018), Arizona (2010-2018), Nevada (2009-2016), Oregon (2014-2018), and Utah (2007-2016). An ED visit was classified as either outpatient or inpatient care directly from the ED. The ED visit record contains service date, age in years, sex, race, zip code, and International Classification of Diseases (ICD) diagnosis codes. Visits before October 1, 2015 used ICD 9^th^ version (ICD-9) and visits afterward used ICD 10^th^ version (ICD-10). The CVDs of interest in this study were acute myocardial infarction (AMI), stroke, HF, atrial fibrillation (AF), and total CVD that was defined as total circulatory disease. We included both the primary and secondary diagnoses of CVDs. The ICD codes used for identification were AMI (ICD-9:410; ICD-10: I21, I22), stroke (ICD-9: 430, 431, 434, 436; ICD-10: I60, I61, I62), HF (ICD-9: 428; ICD-10: I50), AF (ICD-9: 427.3x; ICD-10: I48), and total circulatory diseases (ICD-9: 390-459; ICD-10: I00-I99). This study received approval from the Institutional Review Board (IRB) at Emory University (STUDY00004823), which also granted an exemption from informed consent requirements due to the impracticability of obtaining consent from each individual patient and the minimal risk associated with the study.

### Exposure measures

Daily wildfire smoke PM_2.5_ was estimated using a multi-stage, chemical transport modeling-based framework at a high spatial resolution of 1 km. This model incorporates advanced air quality PM_2.5_ simulations, various satellite remote sensing products, meteorological analyses, land use information, and comprehensive ground-level observations. The total PM_2.5_ model was developed using data from areas impacted by wildfire smoke, while the background PM_2.5_ model was based on data from areas without wildfire smoke. Both models were applied to predict PM_2.5_ concentrations in various scenarios. The difference between total PM_2.5_ and background PM_2.5_ was calculated to isolate the PM_2.5_ contribution from wildfire smoke. Detailed modeling system information has been published elsewhere.^29^ The wildfire smoke, total, and non-smoke PM_2.5_ data were further aggregated to the patients’ zip code level of each day. For all ED records in Nevada, the aggregation was performed by matching the first four digits of the zip codes due to the unavailability of the full zip codes in this state. Additionally, the contribution of smoke PM_2.5_ to total PM_2.5_ were modeled in sensitivity analysis.

### Covariates

In this study, relative humidity, average temperature, and day of the year were accounted for as confounding variables. Daily average temperature was calculated by averaging the minimum and maximum temperatures, and daily relative humidity was estimated using the Magnus formula.^30^ Daymet data on daily minimum and maximum temperatures in degrees Celsius and vapor pressure in pascals, with a resolution of 1 km from 2007 to 2018, were sourced from the Oak Ridge National Laboratory Distributed Active Archive Center for Biogeochemical Dynamics.^31^ The Daymet meteorological data were also aggregated to each patient’s zip code level each day.

### Statistical analysis

We used a case-crossover study design to investigate the associations between short-term smoke PM_2.5_ exposure and ED visits for CVD in the Western United States.^32^ The time-stratified approach was used in the control selection— each ED visit (case) was matched with up to 4 non-event days based on the day of the week in the same calendar month.^33,34^ Because the selection of possible control dates that form a stratum do not depend on the event date, the time-stratified approach is not subject to bias resulting from the time trend.^34^ Additionally, stable characteristics at the individual level (e.g., sex, age, socioeconomic status) are automatically controlled for in the control selection phase. We then performed conditional logistic regression to estimate the associations between smoke PM_2.5_ and total PM_2.5_ with CVD ED visits. We adjusted for non-smoke PM_2.5_ in the model examining the effect of smoke PM_2.5_ because wildfire smoke PM_2.5_ can interact with ambient air pollutants. The model specifications are shown below:

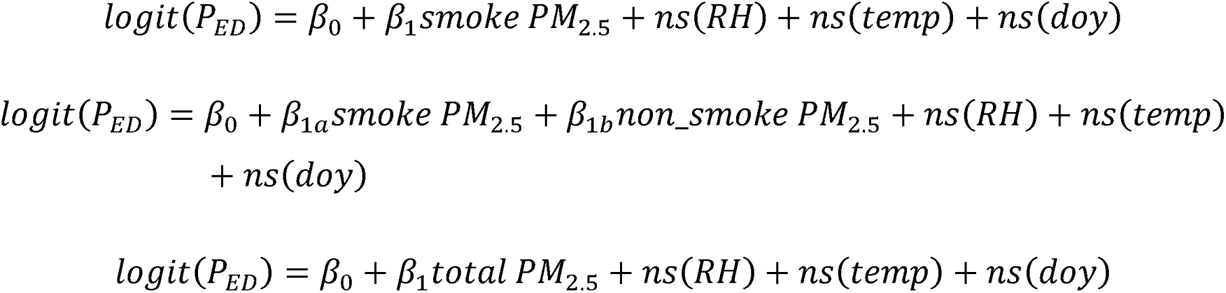

*P_ED_* represents the probability of occurrence of the ED visits for CVD events. For the primary analyses, the outcomes were any primary and secondary diagnosis of the ED visits for CVD events. Relative humidity (RH), average temperature (temp), and day of the year (doy) were modeled using natural splines (ns). The degrees of freedom for the ns function were 3. For wildfire smoke, non-smoke, and total PM_2.5_ concentrations and covariates, we used 1-, 2-, and 3-day averages leading up to and including the day of the ED visit. For sensitivity analysis, we conducted the following: 1) stratified the study population by sex (male and female) and age (<65 years and ≥65 years); 2) examined the effect of the ratio of 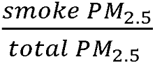 and tested an interaction between the ratio and total PM_2.5_ as following:

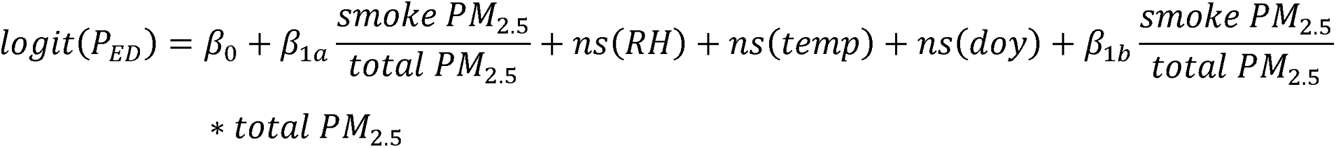

3) estimate the associations between smoke PM_2.5_ and total PM_2.5_ with primary CVD ED visits. Odds ratios (ORs) and their corresponding 95% confidence intervals (95% CIs) were reported. All the analyses were conducted in R 4.2.2 (2022-10-31; available at: https://cran.r-project.org/bin/windows/base/old/4.2.2/).

## Results

In total, 49,759,958 ED visits with a primary or secondary CVD diagnosis were available. Among those, 6,808,839 (14%) were visits for AFs, 1,222,053 (2%) were visits for AMIs, 7,194,474 (14%) were visits for HFs, and 808,396 (2%) were visits for strokes. The average age at the time of the visit was 64 (standard deviation 17.4) years, and 61% of the patients were female. Table 1 shows the characteristics of those who were included in this study with ED visits for primary or secondary diagnoses of CVD. Over the study period from 2007-01-01 to 2018-12-31, the average wildfire smoke, non-smoke, and total PM_2.5_ were 1.11 (IQR 1.29) µg/m^3^, 7.64 (IQR 5.15) µg/m^3^, and 8.77 (IQR 5.77) µg/m^3^, respectively. The distributions of smoke PM_2.5_, total PM_2.5_, non-smoke PM_2.5_, and the ratio of smoke PM_2.5_ and total PM_2.5_ over the study period are shown in Figure 1.

**Figure 1.**
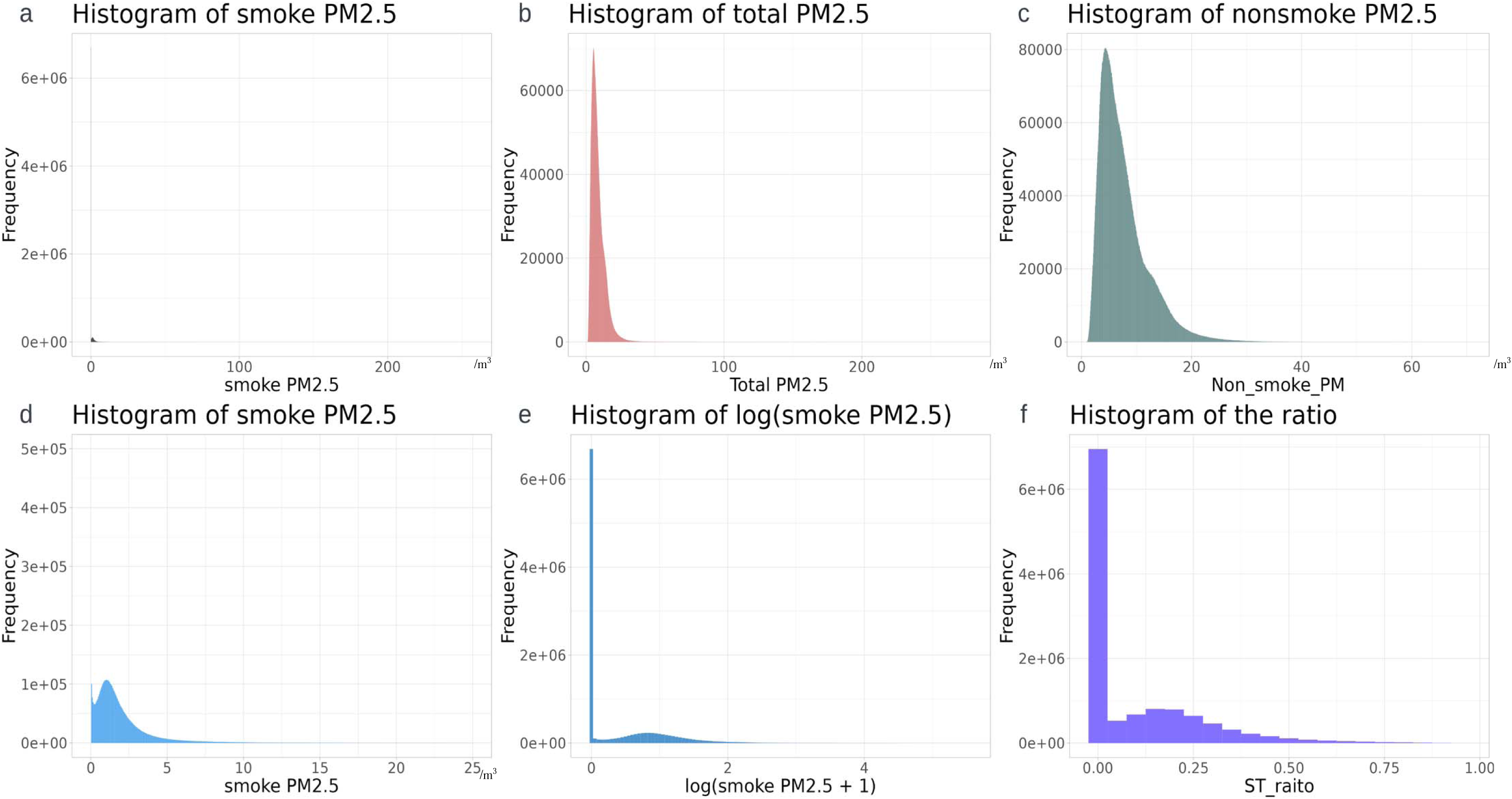
Histograms of smoke PM_2.5_ (a), total smoke PM_2.5_ (b), non-smoke PM_2.5_ (c), smoke PM_2.5_ within 25 /m^3^ and the frequency within 500,000 (d), log (smoke PM_2.5_+1) (e), and the ratio of smoke PM_2.5_/total PM_2.5_ (f). Period 2007-01-01 to 2018-12-31.

**Table 1.**
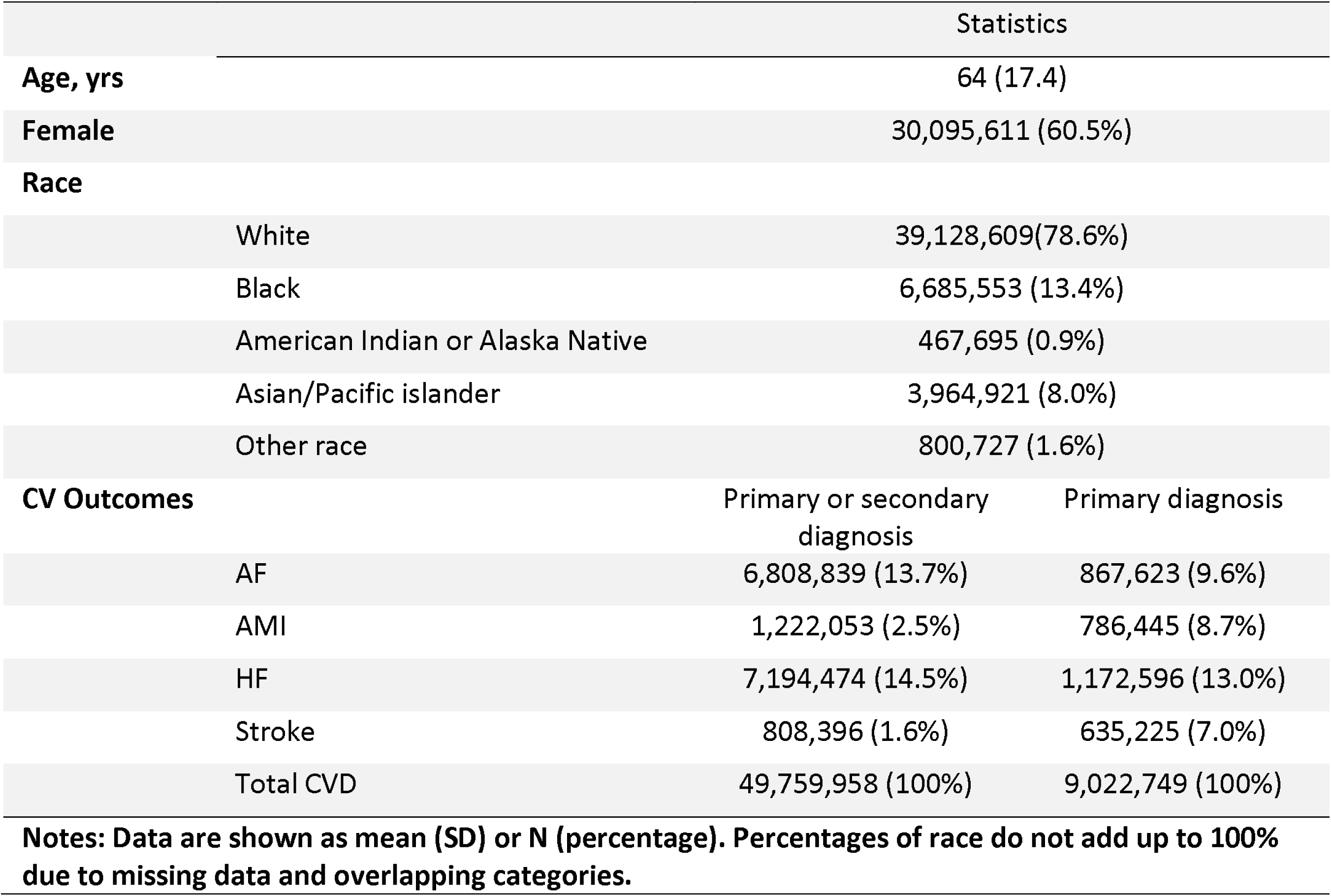
Characteristics of subjects with primary or secondary diagnosis of total CVD (N=49,759, 958)

### Wildfire smoke PM_2.5_ and CVD ED visits in the Western US

During the study period, 10 µg/m^3^ higher smoke PM_2.5_ on the day of the ED visit was associated with a small decrease in the risk of ED visits for AF (OR 0.994, 95% CI 0.991, 0.997), HF (OR 0.995, 95% CI 0.992, 0.998), and CVD (OR 0.997, 95% CI 0.996, 0.998). However, there was no association observed with AMI and stroke. The same results were found between 2-day and 3-day moving average smoke PM_2.5_ and each outcome (Table 2). Further adjusting for non-smoke PM_2.5_ in the model did not change the associations substantively for all CV outcomes. Total PM_2.5_ increased the risk of ED visits for CVDs. A 10 µg/m^3^ higher total PM_2.5_ on the day was associated with a slightly increased risk of ED visits for AF (OR 1.005, 95% CI 1.004, 1.007), AMI (OR 1.010, 95% CI 1.006, 1.015), HF (OR 1.008, 95% CI 1.006, 1.010), and CVD (OR 1.006, 95% CI 1.006, 1.007), but not for stroke (OR 1.001, 95% CI 0.994, 1.007). The 2-day and 3-day moving average total PM_2.5_ showed similar effects on all CV outcomes (Table 2).

**Table 2.** Effect of 1-, 2-, and 3-days smoke PM_2.5_ and total PM_2.5_ on CVD ED visits.

|  |  | AF | AMI | HF | Stroke | CVD |
| --- | --- | --- | --- | --- | --- | --- |
| Smoke PM<sub>2.5</sub> |  |  |  |  |  |  |
| Not adjusting for non-smoke | 1-day | 0.994 (0.991, 0.997) | 0.999 (0.992, 1.006) | 0.995 (0.992, 0.998) | 0.996 (0.985, 1.008) | 0.997 (0.996, 0.998) |
|  | 2-day | 0.992 (0.989, 0.995) | 0.997 (0.990, 1.005) | 0.994 (0.991, 0.997) | 0.996 (0.984, 1.008) | 0.996 (0.993, 0.999) |
|  | 3-day | 0.992 (0.989, 0.995) | 0.998 (0.990, 1.006) | 0.994 (0.991, 0.997) | 0.993 (0.980, 1.006) | 0.996 (0.995, 0.997) |
| Adjusting for non-smoke | 1-day | 0.991 (0.988, 0.994) | 0.997 (0.989, 1.004) | 0.992 (0.989, 0.995) | 0.996 (0.985, 1.007) | 0.995 (0.994, 0.996) |
|  | 2-day | 0.989 (0.986, 0.992) | 0.994 (0.987, 1.002) | 0.990 (0.987, 0.993) | 0.996 (0.984, 1.008) | 0.994 (0.993, 0.995) |
|  | 3-day | 0.989 (0.986, 0.992) | 0.994 (0.986, 1.002) | 0.990 (0.987, 0.993) | 0.993 (0.980, 1.006) | 0.993 (0.992, 0.995) |
| Total PM<sub>2.5</sub> |  |  |  |  |  |  |
|  | 1-day | 1.005 (1.004, 1.007) | 1.010 (1.006, 1.015) | 1.008 (1.006, 1.010) | 1.001 (0.994, 1.007) | 1.006 (1.006, 1.007) |
|  | 2-day | 1.005 (1.003, 1.007) | 1.010 (1.005, 1.010) | 1.008 (1.006, 1.010) | 0.999 (0.991, 1.006) | 1.006 (1.005, 1.007) |
|  | 3-day | 1.005 (1.003, 1.007) | 1.010 (1.005, 1.016) | 1.008 (1.006, 1.010) | 0.995 (0.988, 1.003) | 1.006 (1.005, 1.007) |

Notes: Odds ratios and 95% confidence intervals are presented. AF, atrial fibrillation; AMI, acute myocardial infarction; HF, heart failure; CVD, cardiovascular disease. ORs correspond to every 10 $\mu\text{g}/\text{m}^3$ increase in $\text{PM}_{2.5}$ . All the models were adjusted for daily relative humidity, average temperature, and day of the year. Exposure and covariates were modeled as the value on the day of ED visit, and 2- and 3-day moving averages leading up to and including the day of the ED visit.

### Stratified analysis

The analysis stratified by age (<65 and ≥ 65 years) provided similar results to the main analysis. In both age groups, an increased 1-, 2-, and 3-day smoke resulted in a slight decrease in the risk or no risk change of all CV outcomes. Adjusting for non-smoke PM_2.5_ in addition did not change the results significantly. Higher 1-, 2-, and 3-day total PM_2.5_ were associated with higher risks of ED visits for all CV outcomes (Supplemental Table 1). Sex minimally modified the association between smoke PM_2.5_ and HF, but not for AF, AMI, stroke, and CVD. Similar to the results in the total population, higher total PM_2.5_ was associated with a marginally elevated risk of ED visits for all CV outcomes except stroke among both males and females (Supplemental Table 2).

### The ratio of smoke PM_2.5_ and total PM_2.5_ with CVD ED visits

In the model where the ratio of smoke PM_2.5_ to total PM_2.5_ was modeled as a continuous variable, a 10% increase in the ratio was related to a slightly lower risk of ED visits for AF (1-day OR:0.998, 95% CI 0.997, 0.9997), HF (1-day OR: 0.997, 95% CI 0.996, 0.998), and CVD (1-day OR: 0.999, 95% CI 0.998, 0.999). No significant association was found between the ratio and the risk of ED visits for AMI and stroke. The quartiles of the ratio did not show a remarkable effect on any of the outcomes consistently across models using different days of exposure (Supplemental Table 3).

### Wildfire smoke PM_2.5_ and primary CVD ED visits

A 10 µg/m^3^ higher smoke PM_2.5_ in the model of 1-, 2-, and 3-day was associated with slightly lower risks of HF and total CVD, but not with the risks of AF, AMI, and stroke consistently. Further adjusting for non-smoke did not modify the results. Total PM_2.5_ showed an effect on increasing the risk of all the primary CV endpoints, and the results were only significant for total CVD (Supplemental Table 4).

## Discussion

This study explored the association between short-term PM_2.5_ from wildland fires and ED visits for CVDs in five states in the Western US from 2007 to 2018. With a large sample size of over 49 million, we were able to detect very small effect sizes. Consistent with prior research, total PM_2.5_ was associated with an increased risk of ED visits for CVD.^13,35^ Unexpectedly, though, we found small, but statistically significant, reductions in risk of ED visits for CVD associated with higher exposure to smoke PM_2.5_. The results were not substantially different across different sex and age (<65 and :2 65 years) groups.

Current evidence from epidemiological or environmental exposure studies regarding the links between short-term smoke PM_2.5_ and CVDs is inconsistent, but in general does not support increased risk of CVD with greater short-term exposure to smoke PM_2.5_. A recent review of the impact of wildfires on CV health highlighted that the existing evidence linking CVD with PM_2.5_ and other air pollutants is mixed.^36^ In a US study from 2008 to 2010, increases in PM_2.5_ in days with wildfire smoke exposure (smoke days) were similarly associated with all-cause cardiovascular hospitalizations as similar increases in non-wildfire smoke days.^37^ A similar study in Colorado reported null results regarding the link between smoke PM_2.5_ and any CVD (OR: 0.998, 95% CI 0.984, 1.011) or specific types of CVD including AMI, HF, dysrhythmia, ischemic heart disease and peripheral or cerebrovascular disease, with small effect sizes.^27^ In a study investigating source-apportioned PM_2.5_ and ED visits for CVD in Atlanta, US, biomass burning PM_2.5_ with lag 0 did not present a significant effect on the risk of CVD ED visits; total ambient PM_2.5_ with lag 0 was not associated with most CV outcomes except for ischemic stroke.^38^ During the wildfire period in Canada, no increased risk of physician visits for CVD was observed related to an elevated 2-day PM_2.5_, but in the post-wildfire period, an increased risk of physician visits for HF (11%, 95% CI 3%-21%) and ischemic heart disease (19%, 95% CI 7%-33%) among seniors was found.^39^ It has been reported that the wildfire-related PM_2.5_ in California from 2015 to 2017 was unfavorably associated with a higher risk of out-of-hospital cardiac arrest across multiple lag days (2 day OR: 1.7, 95% CI 1.18, 2.13).^40^ A study in 204 US counties found short-term exposure to ambient PM_2.5_ to be associated with an increased risk of CVD hospital admission rates without statistical significance.^41^ However, the measurements of PM_2.5_ were in three density categories, which reduced statistical power and precision. It is important to note that the cardiovascular associations with PM_2.5_ vary depending on the duration of exposure (short-term vs. long-term), the source of PM_2.5_, and the air pollution regulations in the area where PM_2.5_ levels are high. Furthermore, different study designs, air quality measurements and monitoring, lack of long-term follow-up, and neglect of gaseous species and other hazardous pollutants also contribute to the controversial findings between smoke PM_2.5_ and CVD ED visits.

Several factors may explain the lack of increased CVD risk associated with short-term exposure to smoke PM_2.5_. First, the impact of smoke PM_2.5_ within a short window of exposure might be too subtle to affect acute ED visits for CVD compared to ambient PM_2.5_. While fire smoke is becoming a major contributor of PM_2.5_ pollution in the US, the estimated contribution was up to 25% of total PM_2.5_ in the contiguous US,^11^ and the long-term effect of wildfire smoke PM_2.5_ on CVD might be limited compared with ambient PM_2.5_.^42^ Smoke plumes are often localized and episodic, causing significant variations in PM_2.5_ levels in short durations, while non-smoke PM_2.5_ is more widespread and persistent. The stable temporal and spatial patterns of non-smoke PM_2.5_ might have led to a broader and consistent population impact on CVD risk. In the case that the short-term total PM_2.5_ serves as a proxy for long-term exposure, the observed impact of total PM_2.5_ may be attributed to prolonged exposure. Moreover, the progression of each type of CVD is complex and multifactorial. The precipitating conditions involved in the hypothesized mechanisms of CVD may require a long time to develop, such as arterial occlusion and ischemic necrosis for AMI and stroke,^43^ left ventricular diastolic or systolic dysfunction for HF,^44^ and vulnerable atrial structural and functional substrates for AF.^45^ The risk of acute CVD attributable to several days of exposure to wildfire smoke PM_2.5_ may be diminutive compared to other well-established risk factors. Second, public health advisories and regulations during wildfire seasons and events, and awareness of susceptibility to air pollution may mitigate the adverse health effects of smoke PM_2.5_, as people might stay indoors or seek shelter during wildfires, thereby reducing their true exposure to high levels of smoke PM_2.5_. Moreover, the pattern of behavior change might be different among younger and older adults.^46^ Third, although the case-crossover study design can naturally control for confounding by non-time-dependent factors, time-varying factors and other behaviors subject to change might still confound the associations. For example, some people might stop engaging in outdoor physical activities and stay indoor during the wildfire episode. Changes in behavioral risk factors in days with high smoke PM_2.5_ could even explain the small, but significant, reduction in CVD risk with higher short-term smoke PM_2.5_ exposure. Fourth, the presence of other air pollutants, such as CO, SO_2_, and O_3_, due to wildfires may contribute to the risk of acute CVD, but these were not fully accounted for in this study.

This study has several strengths. This study included a large number of ED visits for CV outcomes across five US states over 12 years, enabling us to detect the small effect of wildfire smoke PM_2.5_. Additionally, the exposure was measured using a high-performance satellite-driven approach with high spatial resolution, offering complete coverage and greater accuracy than monitor-based data. However, several limitations should be acknowledged. First, ED visit data only capture acute CVD events. CVD events requiring long-term medical attention were not included. Second, the exposure and ED visit data were at the zip code level. Misclassification of the patient’s true residential zip code and mismeasurement of the smoke PM_2.5_ exposure may lead to information bias. Third, as mentioned above, residual time-dependent confounding was possible due to the case-crossover study design and unmeasured air pollutants.

## Conclusion

In conclusion, our study examined the relationship between short-term wildfire PM_2.5_ exposure and ED visits for CVD across five Western US states from 2007 to 2018. Although we found statistical significance between smoke PM_2.5_ and total PM_2.5_ with ED visits for some CV outcomes and total CVD, the effect sizes were small and their public health and clinical significance remains uncertain. Further studies with more advanced study designs and more accurate study measures are warranted to investigate the effect of wildfire smoke PM_2.5_ on CV health.

## Data Availability

In accordance with the agreement with each state, the ED visit data is prohibited from sharing to protect health information. The Daymet daily 1 km meteorological data are available in the Oak Ridge National Laboratory data archive (https://doi.org/10.3334/ORNLDAAC/2129). The zip code level wildfire smoke PM2.5 data are available on Figshare (https://doi.org/10.6084/m9.figshare.25016510).

https://doi.org/10.6084/m9.figshare.25016510

## Acknowledgements

We are grateful for the support of the health data sources listed in the following sentence and their contributing hospitals. The ED visits data used to produce this publication were acquired from the Arizona Department of Health Services; California Office of Statewide Planning and Development, now California Department of Health Care Access and Information; Nevada Division of Health Care Financing and Policy (DJCFP), released through the Center for Health Information Analysis (CHIA) of the University of Nevada, Las Vegas; Oregon Healthcare Enterprises, Inc., Apprise Health Insights, a subsidiary of the Oregon Association of Hospitals & Health Systems; and Utah Department of Health, Office of Health Care Statistics (OHCS). The authors thank the ENVISION investigators for their enthusiastic collaboration and effort. The analysis, interpretation, conclusions, and views presented in this paper are solely those of the authors and do not reflect the official positions or conclusions of the listed data sources. The authorization to release this information does not imply any endorsement of the study or its findings by these data sources. The data sources, along with their employees, officers, and agents, do not make any representations, warranties, or guarantees regarding the accuracy, completeness, timeliness, or suitability of the information presented here.

## Disclosures

None.

## Data sharing

In accordance with the agreement with each state, the ED visit data is prohibited from sharing to protect health information. The Daymet daily 1 km meteorological data are available in the Oak Ridge National Laboratory data archive (https://doi.org/10.3334/ORNLDAAC/2129). The zip code level wildfire smoke PM_2.5_ data are available on Figshare (https://doi.org/10.6084/m9.figshare.25016510).

## Funding

This work was supported by the National Institutes of Environmental Health Services under Award Number 1R01ES032140 (Liu) and R01ES027892 (Chang), and the American Heart Association under Award Number 23PRE1020888 (Li). The content is solely the responsibility of the authors and does not necessarily represent the official views of the National Institutes of Health.

---

**Supplemental Table 1.** Effect of 1-, 2-, and 3-days smoke PM_2.5_ and total PM_2.5_ on CVD ED visits among those < 65 and ≥ 65 years.

|  |  | AF | AMI | HF | Stroke | CVD |
| --- | --- | --- | --- | --- | --- | --- |
| <65 years |  |  |  |  |  |  |
| Smoke PM<sub>2.5</sub> |  |  |  |  |  |  |
| Not adjusting for non-smoke | 1-day | 0.996 (0.989, 1.002) | 0.993 (0.981, 1.005) | 0.999 (0.994, 1.003) | 0.981 (0.963, 1.000) | 0.998 (0.996, 1.000) |
|  | 2-day | 0.994 (0.987, 1.001) | 0.992 (0.979, 1.005) | 0.998 (0.992, 1.003) | 0.987 (0.968, 1.010) | 0.998 (0.996, 0.999) |
|  | 3-day | 0.994 (0.986, 1.001) | 0.993 (0.979, 1.007) | 0.998 (0.992, 1.004) | 0.983 (0.963, 1.010) | 0.997 (0.995, 0.999) |
| Adjusting for non-smoke | 1-day | 0.994 (0.988, 1.001) | 0.991 (0.979, 1.004) | 0.997 (0.992, 1.002) | 0.981 (0.963, 1.000) | 0.996 (0.995, 0.998) |
|  | 2-day | 0.992 (0.985, 0.999) | 0.990 (0.977, 1.004) | 0.995 (0.990, 1.001) | 0.987 (0.968, 1.010) | 0.996 (0.994, 0.998) |
|  | 3-day | 0.991 (0.984, 0.999) | 0.991 (0.977, 1.005) | 0.995 (0.990, 1.001) | 0.984 (0.963, 1.010) | 0.995 (0.993, 0.997) |
| Total PM<sub>2.5</sub> |  |  |  |  |  |  |
|  | 1-day | 1.003 (0.998, 1.007) | 1.004 (0.996, 1.012) | 1.006 (1.003, 1.009) | 0.995 (0.984, 1.010) | 1.005 (1.004, 1.010) |
|  | 2-day | 1.002 (0.998, 1.007) | 1.002 (0.993, 1.010) | 1.006 (1.002, 1.009) | 0.993 (0.981, 1.000) | 1.005 (1.004, 1.010) |
|  | 3-day | 1.002 (0.997, 1.007) | 1.000 (0.991, 1.010) | 1.006 (1.002, 1.010) | 0.990 (0.977, 1.000) | 1.005 (1.003, 1.010) |
| ≥65 years |  |  |  |  |  |  |
| Smoke PM<sub>2.5</sub> |  |  |  |  |  |  |
| Not adjusting for non-smoke | 1-day | 0.993 (0.990, 0.997) | 1.002 (0.993, 1.011) | 0.994 (0.990, 0.997) | 1.005 (0.991, 1.019) | 0.996 (0.994, 0.998) |
|  | 2-day | 0.992 (0.988, 0.995) | 1.000 (0.991, 1.009) | 0.992 (0.989, 0.996) | 1.001 (0.986, 1.016) | 0.995 (0.994, 0.997) |
|  | 3-day | 0.992 (0.988, 0.996) | 1.000 (0.990, 1.010) | 0.992 (0.988, 0.996) | 0.997 (0.981, 1.014) | 0.995 (0.993, 0.997) |
| Adjusting for non-smoke | 1-day | 0.991 (0.988, 0.994) | 0.999 (0.990, 1.010) | 0.990 (0.987, 0.994) | 1.004 (0.990, 1.018) | 0.994 (0.992, 0.995) |
|  | 2-day | 0.989 (0.985, 0.992) | 0.996 (0.987, 1.005) | 0.988 (0.985, 0.992) | 1.001 (0.986, 1.016) | 0.993 (0.991, 0.994) |
|  | 3-day | 0.989 (0.985, 0.992) | 0.995 (0.985, 1.005) | 0.988 (0.984, 0.991) | 0.998 (0.982, 1.014) | 0.992 (0.990, 0.994) |
| Total PM<sub>2.5</sub> |  |  |  |  |  |  |
|  | 1-day | 1.005 (1.003, 1.008) | 1.013 (1.008, 1.019) | 1.008 (1.006, 1.011) | 1.005 (0.997, 1.013) | 1.007 (1.006, 1.008) |
|  | 2-day | 1.005 (1.002, 1.007) | 1.014 (1.007, 1.020) | 1.008 (1.006, 1.010) | 1.001 (0.992, 1.010) | 1.007 (1.005, 1.008) |
|  | 3-day | 1.005 (1.002, 1.007) | 1.015 (1.008, 1.020) | 1.008 (1.006, 1.011) | 0.997 (0.988, 1.007) | 1.006 (1.005, 1.007) |
| Notes: Odds ratios and 95% confidence intervals are presented. AF, atrial fibrillation; AMI, acute myocardial infarction; HF, heart failure; CVD, cardiovascular disease. ORs correspond to every 10 μg/m<sup>3</sup> increase in PM<sub>2.5</sub>. All the models were adjusted for daily relative humidity, average temperature, and day of the year. Exposure and covariates were modeled as the value on the day of ED visit, and 2- and 3-day moving averages leading up to and including the day of the ED visit. |  |  |  |  |  |  |

**Supplemental Table 2.** Effect of 1-, 2-, and 3-days smoke PM_2.5_ and total PM_2.5_ on CVD ED visits among different sex groups.

|  |  | AF | AMI | HF | Stroke | CVD |
| --- | --- | --- | --- | --- | --- | --- |
| **Male** |  |  |  |  |  |  |
| **Smoke PM<sub>2.5</sub>** |  |  |  |  |  |  |
| *Not adjusting for non-smoke* | 1-day | 0.992 (0.987, 0.997) | 0.996 (0.983, 1.010) | 0.995 (0.990, 1.000) | 0.991 (0.972, 1.009) | 0.997 (0.995, 0.999) |
|  | 2-day | 0.989 (0.984, 0.995) | 0.993 (0.978, 1.008) | 0.992 (0.987, 0.998) | 0.987 (0.967, 1.007) | 0.996 (0.993, 0.998) |
|  | 3-day | 0.989 (0.984, 0.995) | 0.994 (0.979, 1.010) | 0.991 (0.986, 0.997) | 0.842 (0.962, 1.004) | 0.995 (0.992, 0.997) |
| *Adjusting for non-smoke* | 1-day | 0.990 (0.985, 0.995) | 0.994 (0.981, 1.008) | 0.992 (0.987, 0.998) | 0.990 (0.971, 1.009) | 0.995 (0.993, 0.997) |
|  | 2-day | 0.987 (0.981, 0.992) | 0.991 (0.976, 1.006) | 0.989 (0.984, 0.995) | 0.987 (0.967, 1.008) | 0.993 (0.991, 0.995) |
|  | 3-day | 0.987 (0.981, 0.992) | 0.992 (0.976, 1.008) | 0.988 (0.982, 0.994) | 0.983 (0.962, 1.005) | 0.992 (0.990, 0.994) |
| **Total PM<sub>2.5</sub>** |  |  |  |  |  |  |
|  | 1-day | 1.003 (1.000, 1.006) | 1.008 (1.000, 1.015) | 1.006 (1.003, 1.009) | 1.000 (0.990, 1.010) | 1.006 (1.005, 1.007) |
|  | 2-day | 1.004 (1.000, 1.007) | 1.007 (0.998, 1.020) | 1.007 (1.004, 1.010) | 0.994 (0.983, 1.005) | 1.006 (1.004, 1.007) |
|  | 3-day | 1.004 (1.000, 1.007) | 1.007 (0.998, 1.020) | 1.007 (1.000, 1.011) | 0.990 (0.978, 1.002) | 1.005 (1.004, 1.007) |
| **Female** |  |  |  |  |  |  |
| **Smoke PM<sub>2.5</sub>** |  |  |  |  |  |  |
| *Not adjusting for non-smoke* | 1-day | 0.995 (0.990, 1.000) | 1.012 (0.996, 1.028) | 0.999 (0.994, 1.004) | 0.992 (0.974, 1.010) | 0.998 (0.996, 0.999) |
|  | 2-day | 0.994 (0.988, 0.999) | 1.012 (0.995, 1.030) | 0.999 (0.993, 1.004) | 0.993 (0.973, 1.013) | 0.997 (0.996, 0.999) |
|  | 3-day | 0.993 (0.988, 0.990) | 1.014 (0.995, 1.033) | 0.998 (0.993, 1.004) | 0.989 (0.968, 1.011) | 0.997 (0.995, 0.990) |
| *Adjusting for non-smoke* | 1-day | 0.993 (0.988, 0.998) | 1.011 (0.994, 1.027) | 0.996 (0.991, 1.001) | 0.991 (0.973, 1.010) | 0.996 (0.994, 0.998) |
|  | 2-day | 0.991 (0.986, 0.997) | 1.011 (0.993, 1.028) | 0.995 (0.990, 1.000) | 0.992 (0.972, 1.012) | 0.995 (0.993, 0.997) |
|  | 3-day | 0.991 (0.985, 0.997) | 1.013 (0.994, 1.032) | 0.994 (0.989, 1.000) | 0.989 (0.967, 1.010) | 0.994 (0.992, 0.996) |
| **Total PM<sub>2.5</sub>** |  |  |  |  |  |  |
|  | 1-day | 1.007 (1.003, 1.010) | 1.010 (1.001, 1.019) | 1.012 (1.009, 1.015) | 1.001 (0.992, 1.011) | 1.007 (1.005, 1.008) |
|  | 2-day | 1.006 (1.003, 1.009) | 1.011 (1.001, 1.021) | 0.999 (0.993, 1.004) | 1.002 (0.992, 1.013) | 1.007 (1.006, 1.008) |
|  | 3-day | 1.006 (1.002, 1.009) | 1.012 (1.001, 1.023) | 1.012 (1.009, 1.016) | 1.000 (0.988, 1.011) | 1.006 (1.005, 1.008) |

Notes: Odds ratios and 95% confidence intervals are presented. AF, atrial fibrillation; AMI, acute myocardial infarction; HF, heart failure; CVD, cardiovascular disease. ORs correspond to every 10 µg/m<sup>3</sup> increase in PM<sub>2.5</sub>. All the models were adjusted for daily relative humidity, average temperature, and day of the year. Exposure was modeled as the PM<sub>2.5</sub> on the day of ED visit, and 2- and 3-day moving averages of the PM<sub>2.5</sub> leading up to and including the day of the ED visit.

**Supplemental Table 3.**
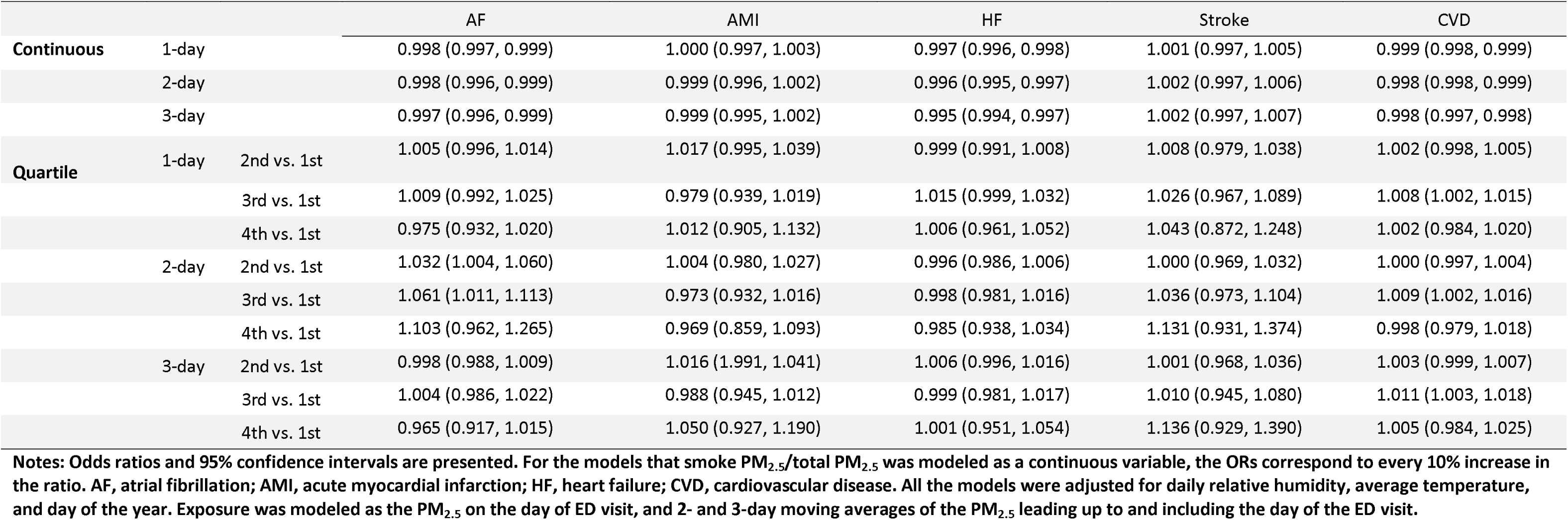
Effect of 1 day, 2-day, and 3-day average ratio of smoke PM_2.5_ and total PM_2.5_ on CVD ED visits.

|  |  |  | AF | AMI | HF | Stroke | CVD |
| --- | --- | --- | --- | --- | --- | --- | --- |
| **Continuous** | 1-day |  | 0.998 (0.997, 0.999) | 1.000 (0.997, 1.003) | 0.997 (0.996, 0.998) | 1.001 (0.997, 1.005) | 0.999 (0.998, 0.999) |
|  | 2-day |  | 0.998 (0.996, 0.999) | 0.999 (0.996, 1.002) | 0.996 (0.995, 0.997) | 1.002 (0.997, 1.006) | 0.998 (0.998, 0.999) |
|  | 3-day |  | 0.997 (0.996, 0.999) | 0.999 (0.995, 1.002) | 0.995 (0.994, 0.997) | 1.002 (0.997, 1.007) | 0.998 (0.997, 0.998) |
| **Quartile** | 1-day | 2nd vs. 1st | 1.005 (0.996, 1.014) | 1.017 (0.995, 1.039) | 0.999 (0.991, 1.008) | 1.008 (0.979, 1.038) | 1.002 (0.998, 1.005) |
|  |  | 3rd vs. 1st | 1.009 (0.992, 1.025) | 0.979 (0.939, 1.019) | 1.015 (0.999, 1.032) | 1.026 (0.967, 1.089) | 1.008 (1.002, 1.015) |
|  |  | 4th vs. 1st | 0.975 (0.932, 1.020) | 1.012 (0.905, 1.132) | 1.006 (0.961, 1.052) | 1.043 (0.872, 1.248) | 1.002 (0.984, 1.020) |
|  | 2-day | 2nd vs. 1st | 1.032 (1.004, 1.060) | 1.004 (0.980, 1.027) | 0.996 (0.986, 1.006) | 1.000 (0.969, 1.032) | 1.000 (0.997, 1.004) |
|  |  | 3rd vs. 1st | 1.061 (1.011, 1.113) | 0.973 (0.932, 1.016) | 0.998 (0.981, 1.016) | 1.036 (0.973, 1.104) | 1.009 (1.002, 1.016) |
|  |  | 4th vs. 1st | 1.103 (0.962, 1.265) | 0.969 (0.859, 1.093) | 0.985 (0.938, 1.034) | 1.131 (0.931, 1.374) | 0.998 (0.979, 1.018) |
|  | 3-day | 2nd vs. 1st | 0.998 (0.988, 1.009) | 1.016 (1.991, 1.041) | 1.006 (0.996, 1.016) | 1.001 (0.968, 1.036) | 1.003 (0.999, 1.007) |
|  |  | 3rd vs. 1st | 1.004 (0.986, 1.022) | 0.988 (0.945, 1.012) | 0.999 (0.981, 1.017) | 1.010 (0.945, 1.080) | 1.011 (1.003, 1.018) |
|  |  | 4th vs. 1st | 0.965 (0.917, 1.015) | 1.050 (0.927, 1.190) | 1.001 (0.951, 1.054) | 1.136 (0.929, 1.390) | 1.005 (0.984, 1.025) |

**Notes:** Odds ratios and 95% confidence intervals are presented. For the models that smoke PM<sub>2.5</sub>/total PM<sub>2.5</sub> was modeled as a continuous variable, the ORs correspond to every 10% increase in the ratio. AF, atrial fibrillation; AMI, acute myocardial infarction; HF, heart failure; CVD, cardiovascular disease. All the models were adjusted for daily relative humidity, average temperature, and day of the year. Exposure was modeled as the PM<sub>2.5</sub> on the day of ED visit, and 2- and 3-day moving averages of the PM<sub>2.5</sub> leading up to and including the day of the ED visit.

**Supplemental Table 4.**
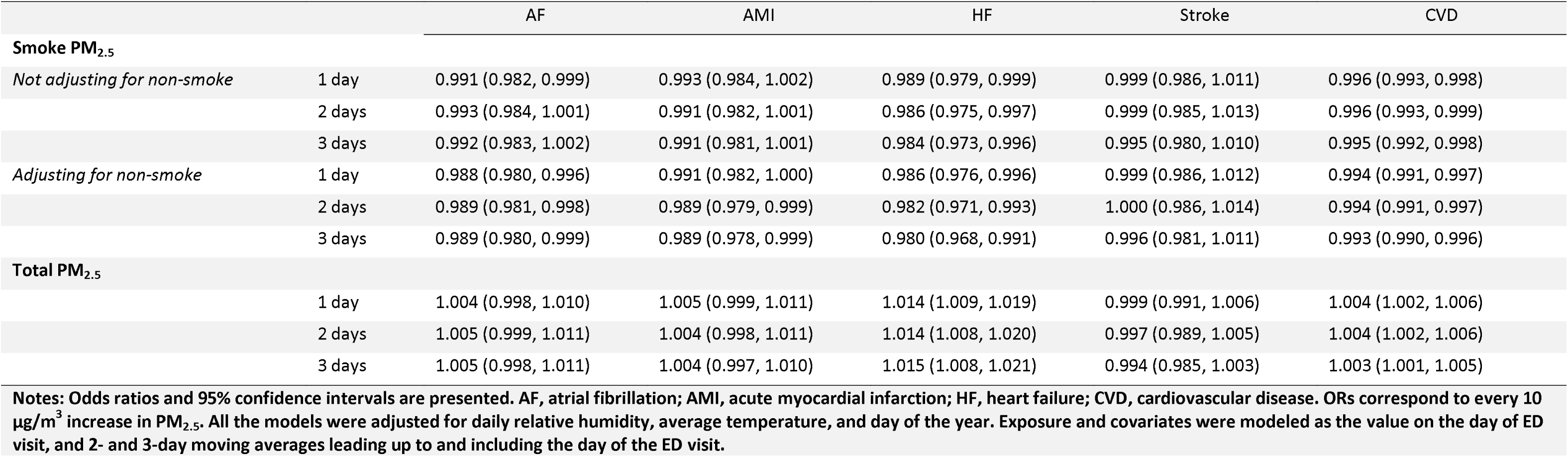
Effect of 1 day, 2-day, and 3-day smoke PM2.5 and total PM2.5 on primary CVD ED visits.

## Notes

### Competing Interest Statement

The authors have declared no competing interest.

### Author Declarations

This study received approval from the Institutional Review Board (IRB) at Emory University (STUDY00004823), which also granted an exemption from informed consent requirements due to the impracticability of obtaining consent from each individual patient and the minimal risk associated with the study.

### Summary of Updates

Added ORCID for all authors; revised funding information.

